# Populism Versus Public Health: Lessons from Iran’s COVID-19 Crisis and the Global Cost of Scientific Misinformation

**DOI:** 10.1101/2024.12.28.24319727

**Authors:** Nima Taefehshokr, Brielle Martens, Iman Beheshti, Rene P Zahedi, Marek J Los, Jason Kindrachuk, Saeid Ghavami

**Affiliations:** Department of Human Anatomy and Cell Science, Max Rady College of Medicine, Rady Faculty of Health Sciences, University of Manitoba, Winnipeg, MB R3T 2N2, Canada; Department of Medical Microbiology and Infectious Diseases, University of Manitoba, Winnipeg, Canada; Department of Human Anatomy and Cell Science, University of Manitoba, Winnipeg, MB, Canada; Department of Biochemistry and Medical Genetics, University of Manitoba, Winnipeg, Manitoba R3E 0J9, Canada; Biotechnology Center, Silesian University of Technology, Gliwice, Poland; Faculty of Medicine, Rolna 43, Katowice, Poland; Paul Albrechtsen Research Institute, CancerCare Manitoba, Winnipeg, MB, Canada; Department of Human Anatomy and Cell Science, Max Rady College of Medicine, Rady Faculty of Health Sciences, University of Manitoba, Winnipeg, MB, Canada

**Keywords:** Science-Related Populism, Public Health Misinformation, COVID-19 Policy Impacts, Global Health Consequences, Iranian COVID-19 Response, Machine Learning

## Abstract

This article explores the impact of science-related populism on public health during the COVID-19 pandemic, with a focus on the situation in Iran. Our analysis of publicly available data from The Economist and the World Health Organization demonstrates the consequences of delayed vaccination campaigns, showing a clear correlation between delayed vaccine introduction and increased excess mortality rates. This trend is particularly pronounced in Iran, where political resistance to Western vaccine imports and the promotion of unproven medical technologies exacerbated the public health crisis. Additionally, our time-series analysis links significant surges in COVID-19 deaths to government related events and decisions that likely enhanced virus transmission, indicating direct public health repercussions from delayed pandemic responses. These results highlight the global security threat posed by science-related populism, where political agendas undermine scientific integrity and public health. The findings advocate for an urgent global commitment to uphold scientific evidence as the cornerstone of health policy, emphasizing the necessity of combating misinformation to ensure timely and effective public health measures.

## Introduction

Populist leaders frequently portray themselves as defenders of “the people” against so-called elites, including scientists and experts. This can create a cultural climate in which scientific expertise is devalued, with populist leaders instead offering oversimplified solutions to complex problems [1, 2]. We observed this globally during the COVID-19 pandemic [3-5], and unfortunately current events in the United States have taken populism to an extreme few could have predicted [6]. Science-related populism is a rising complex global challenge that undermines public trust in science and interferes with progress on tackling global crises [7]. Although often viewed as a national issue, science-related populism in one country can have far-reaching global consequences [8]. The current US situation notwithstanding, this analysis explores science-related populism in Iran during the COVID-19 pandemic and its resulting broader implications for global health [3, 9]. We argue that the promotion by political leaders of pseudoscience and ideological rhetoric over science-based evidence poses a significant threat to global public health.

In Iran, the COVID-19 crisis was marked by the overshadowing of evidence-based decision-making by populist political interests. The problem became evident when the Iranian government promoted the “Mostaan 110” device, which was purported, with no evidence, to detect COVID-19 from a distance. Despite being debunked by the Iranian Physics Society as pseudoscientific [10, 11], the device continued to be endorsed by political leaders. This political promotion of pseudoscience sowed confusion and mistrust within Iran and internationally [11]. Iran’s political stance during the COVID-19 pandemic significantly worsened the public health crisis by delaying vaccination efforts. Hardline leaders prioritized political agendas over scientific recommendations, banning vaccine imports from the United States and United Kingdom, labeling them as “biological weapons” [12]. Vaccine shortages delayed the start of Iran’s national vaccination campaign [12]. This situation was influenced by challenges with importing vaccines from abroad. The country’s former health minister, Dr. Bahram Eynollahi, along with 2,500 other doctors, expressed concerns about importing vaccines in an open letter to the Iranian president in February 2021 [12]. The letter instead promoted the domestic vaccines Noora and SpikoGen as alternatives. The clinical trial of the Noora vaccine has since been retracted [13], and a recent report showed weak immunogenicity compared to other vaccines [14]. The rejection of imported vaccines delayed the national vaccination campaign, leaving millions of Iranians vulnerable.

## Vaccine delay led to elevated excess deaths

To demonstrate the impact of vaccination delay in Iran, we used publicly available data from The Economist’s excess deaths model and the World Health Organization (WHO) [15-17]. Data was analyzed in RStudio (v.2024.09.1+394) using the dplyr (v.1.1.4) and lubridate (v.1.9.4) packages [18, 19]. The ggplot2 (v.3.5.1) and cowplot (v.1.1.1) packages were used to generate figures [20, 21].

Figure 1 shows the relationship between cumulative COVID-19 vaccinations and estimated excess deaths per 100,000 across multiple countries. This analysis highlights a stark contrast not only between early and delayed vaccine introduction but also the impact of the degree of vaccine uptake. Countries such as Bahrain and Canada demonstrated early vaccination starts, reflected in lower excess deaths. The United States, which introduced vaccines at the same time as Canada, did not achieve the same level of vaccine uptake, resulting in a much higher excess mortality. Some nations, including Iran and Montenegro, did not introduce vaccines until well into February of 2024, with accompanying low vaccine uptake and resulting in elevated excess mortality. In the case of Iran, vaccine imports were delayed due to political decisions, and the country’s vaccination campaign lagged, marked by late vaccine introduction and limited uptake, correlating with a steep rise in excess deaths. Compared to countries like Canada and Bahrain, which rapidly implemented vaccination programs, Iran’s policy-driven delay clearly amplified the nation’s death toll.

**Figure 1.**
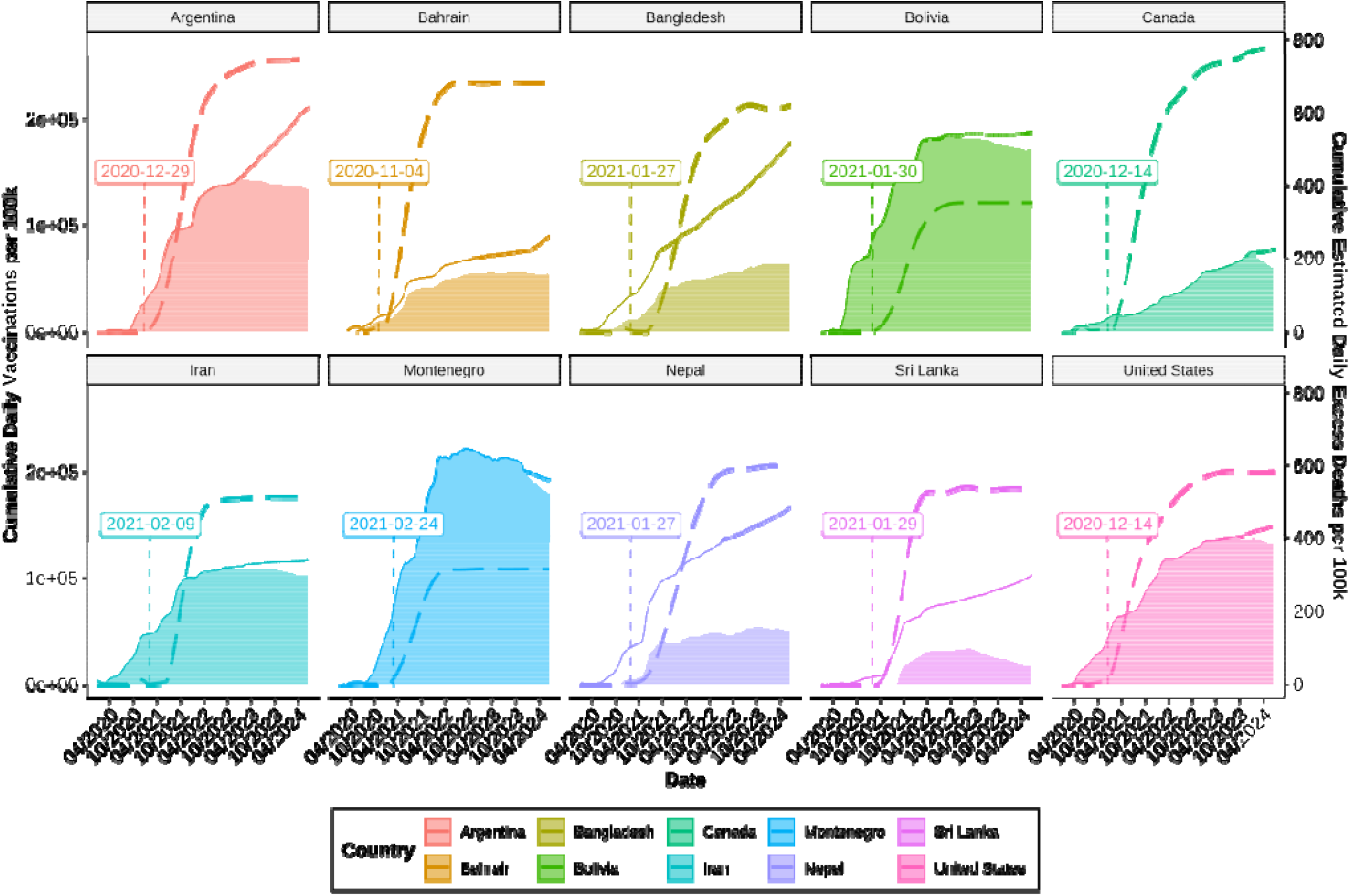
Cumulative vaccinations and estimated excess deaths due to COVID-19 per 100,000 population over time. The date of the first COVID-19 vaccine introduction is shown for each country. Dashed line corresponds to cumulative daily vaccinations. The area plot corresponds to estimated excess deaths, the darker and lighter shaded regions represent the lower and upper 95% confidence intervals, respectively. Data was obtained from the World Health Organization and The Economist’s excess deaths model [15-17].

## Vaccine delay correlates inversely with vaccine-averted COVID-19 deaths

We obtained additional data from the WHO and Ghafari et al. [16, 17, 22] for the Eastern Mediterranean WHO region. Data was analyzed using RStudio (v.2024.09.1+394). The ggplot2 (v.3.5.1) package was used to generate figures [20]. Vaccine delay was calculated by taking the difference between the date of first vaccine introduction for each country and their WHO region. The rstatix (v.0.7.2) package [23] was used to conduct a regression test to determine whether there was a significant relationship between the number of deaths averted by vaccination and vaccine delay. Our results are plotted in Figure 2.

**Figure 2.**
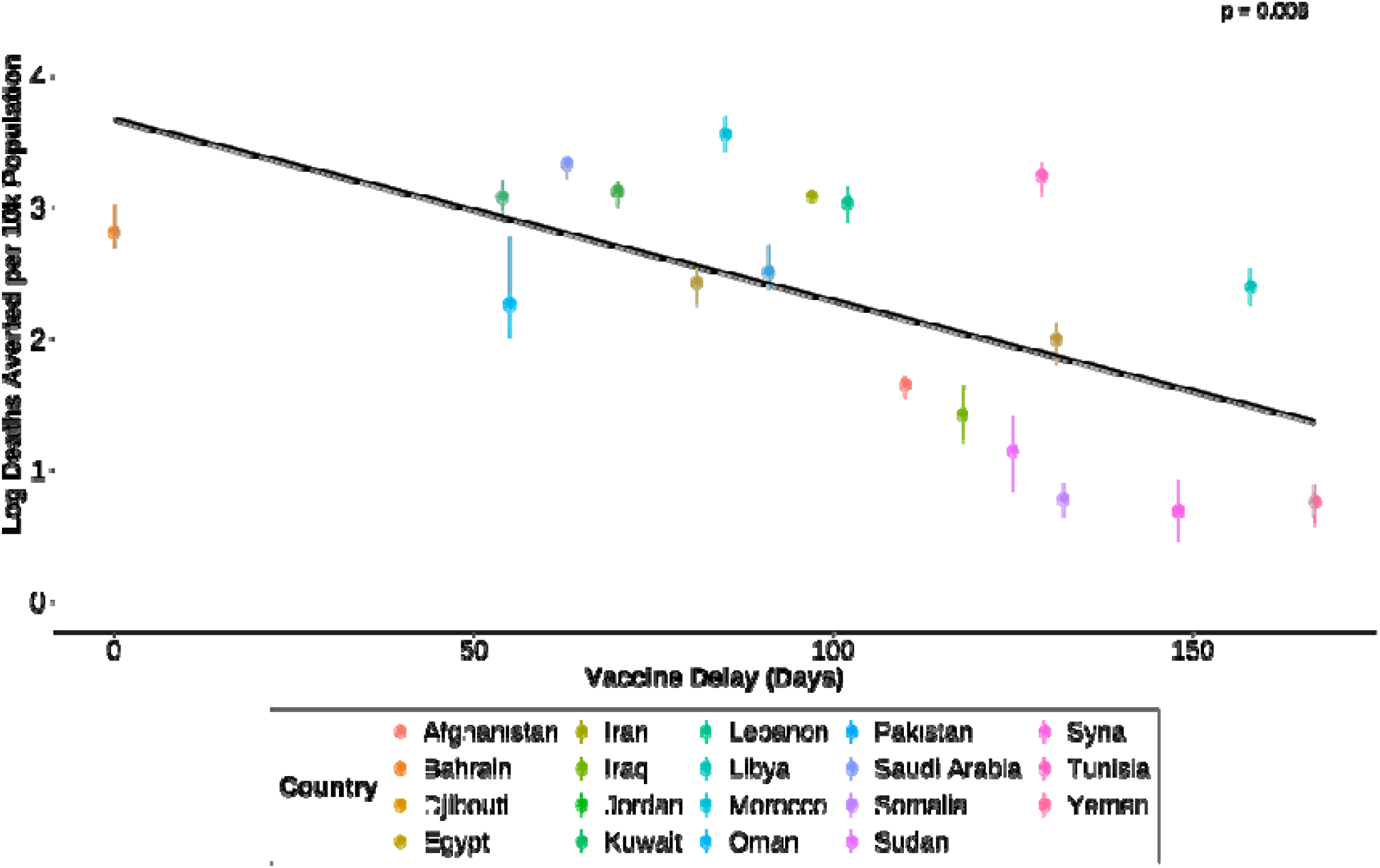
Linear association between vaccine delay and the number of deaths averted by COVID-19 vaccines per 10,000 population across countries in the WHO Eastern Mediterranean Region. Each point represents a country. Vertical bars represent 95% confidence intervals. A linear regression model was applied; the regression line is shown in black with the shaded area representing the 95% confidence band. The p-value tests the null hypothesis that the slope of the regression line is equal to zero. Data sources: WHO and Ghafari et al.[16, 17, 22].

We found a significant inverse relationship. The figure highlights Iran’s notably delayed vaccine rollout, which resulted in substantially fewer averted deaths compared to regional counterparts. In contrast, countries such as Bahrain and the United Arab Emirates, which implemented swift vaccination campaigns, achieved far better outcomes in terms of lives saved. This finding underscores the dire consequences of delayed vaccine access and the importance of prompt, science-driven public health responses to mitigate large-scale health crises.

## Impact of scientific populism on waves of COVID-19 deaths reported in Iran

Using previously published data [24], we analyzed waves of COVID-19 deaths from 2020 to 2022 to further evaluate the impact of scientific populism in Iran’s experience of the COVID-19 pandemic. In this time period, Iran experienced five waves of COVID-19 deaths; these are charted in Figure 3A. The first wave occurred shortly after the parliamentary election held on 21 February 2020. The Iranian government downplayed the seriousness of the virus at the beginning of 2020 in order to maximize voter turnout. Then in August 2020, a second wave occurred after the government widely encouraged public participation in mourning ceremonies, processions, and gatherings associated with the sacred months of Muharram and Safar, including the observation of Ashura through mourning gatherings and processions. A third wave occurred during the last quarter of 2020. (Figure 3A). After a period of low reported numbers of infections in the first quarter of 2021, a sudden spike in infections and deaths constituted a fourth wave in April 2021. The fifth wave, which saw the highest number of deaths, coincided with Moharram and Safar in 2021 (Figure 3A).

**Figure 3A:**
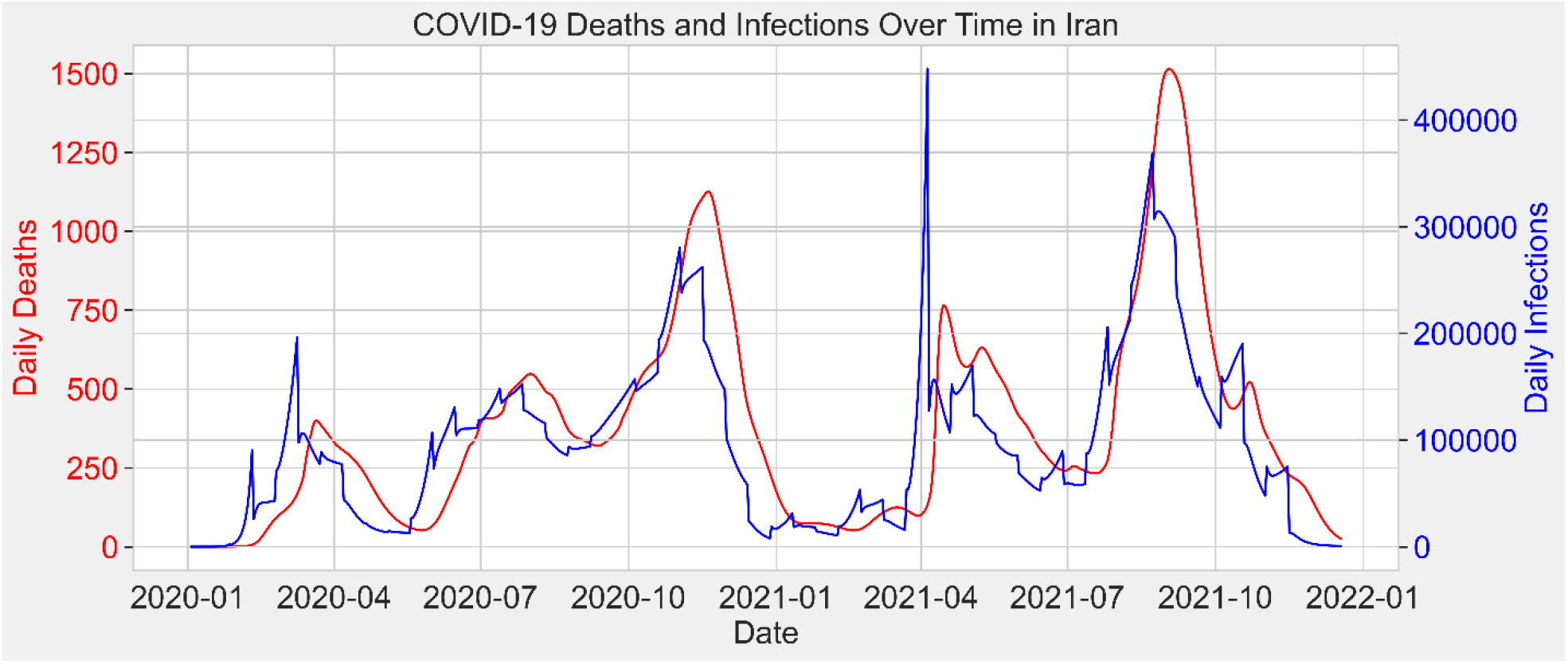
Daily distribution of COVID-19 infections and deaths in Iran from January 2020 to January 2022, based on data presented in [24]

**Figure 3B:**
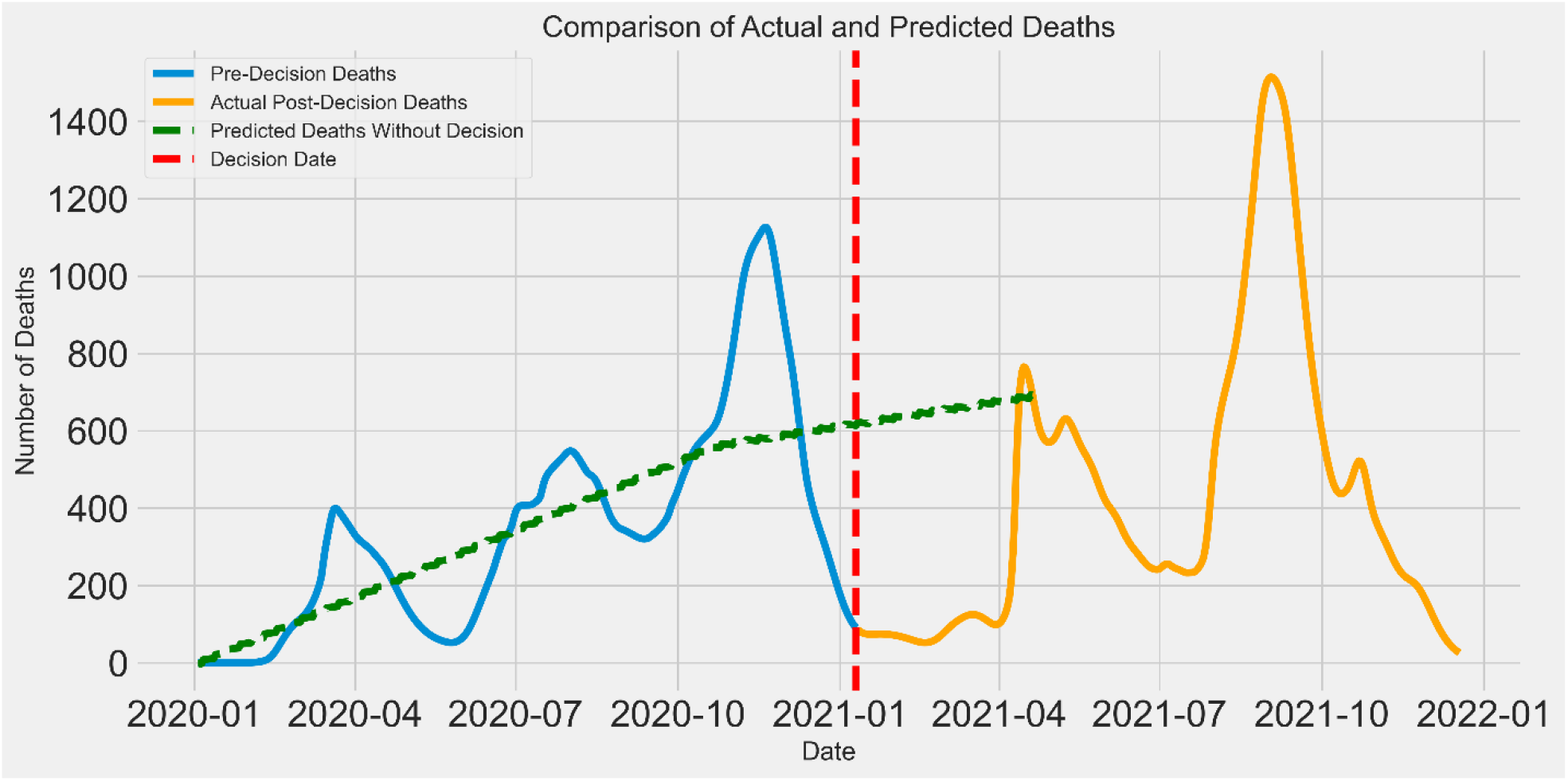
Time series comparison of reported COVID-19 deaths before and after the government’s political announcement in January 2021, overlaid with the predicted number of deaths had no policy change occurred. The predictions were generated using **Prophet**, a validated forecasting algorithm that models time series data with components for trend, seasonality, and holiday effects (https://facebook.github.io/prophet/). The model was trained on data **prior to the policy decision**, and forecasts represent the expected trajectory **in the absence of that decision**.

Based on the open letter discussed in the Introduction above, in the spring of 2021 the Supreme Leader took a decision against the import of foreign-made vaccines from the United States and the United Kingdom in favor of the Iranian-made Noora and SpikeGen vaccines. In the time surrounding this entirely political decision – which was based entirely in scientific populism and not at all in scientific evidence – the Iranian government reported a dramatic decline in the number of deaths and infections, and available data shows that this trend persisted until the end of March 2021 (Figure 3A). Many experts have questioned the accuracy of these reports, considering them to be manipulated or falsified [25, 26].

We performed our own time series prediction using Prophet, a validated tool developed by Facebook. Prophet accounts for trends, seasonality, and holidays, making it useful for predicting data that displays such patterns, including COVID-19 cases (for more details, see https://facebook.github.io/prophet/) [27]. We trained the Prophet model using reported data from January 2020 to January 2021 [24] (https://github.com/mrc-ide/covid-vaccine-impact-orderly) and predicted the pattern of daily deaths for 100 days after the political decision in January 2021 (Figure 3B). In contrast to the reported data, our model predicted a mean increase in deaths during the 100 days following this decision. While this discrepancy raises questions about the consistency of reported mortality trends, we acknowledge that multiple factors—including potential model limitations, delayed or incomplete reporting, or genuine epidemiological shifts— could account for the divergence. Nevertheless, given prior reports questioning the accuracy of official statistics in this context [26], the divergence merits further scrutiny.

Regardless, the decision to reject imported vaccines delayed Iran’s vaccination efforts, with direct public health consequences contributing to an estimated more 50,000 deaths [22]. As occurred with many other countries, the elevated fatality rate affected patients of advanced average (mean age: 65.67 □ ± □15.23) with a high prevalence of pre-existing conditions [28], However, Iran as a nation had set its own precent having experienced delays relative to other countries in launching a previous national immunization program in 1984 [29], Overall, the population likely suffers from insufficient cross-immunity in addition the detrimental effects of sanctions on Iran’s economy and healthcare system functionality [30].

## Dangerous consequences of a populist agenda

The Iranian case illustrates the dangerous consequences of allowing populist agendas to distort science. But this is not an isolated phenomenon—it is a global issue that threatens the foundations of public health and scientific integrity. In today’s interconnected world, neither viruses nor misinformation respect geographical boundaries. Misinformation spreads rapidly on social media, further amplifying pseudoscientific claims and making it more difficult for health authorities to implement effective public health measures. Delayed vaccination uptake in one country increases the likelihood that new vaccine-evading variants of concern will emerge [31, 32]. Although vaccine hesitancy and populist rhetoric during the pandemic were similarly observed in countries such as Brazil and the United States [33, 34], it was in Iran that the science-related populism took hold at the highest level of decision-making power, impacting the entire country’s population. The danger of allowing pseudoscience to flourish in environments where political narratives overshadow scientific scrutiny is undeniable, with far-reaching effects. Unfortunately, this is becoming all to clear under the current US presidency of Donald Trump, who administration has set scientific progress back in the country by decades, with National Institutes of Health and the Centers for Disease control canceling billions in funding for COVID-related research and pandemic preparedness [35].

Now, more than ever, the global scientific community cannot afford to be passive observers. Collective action is needed to ensure that scientific evidence, not political rhetoric, guides global health decisions. The international scientific community must actively engage in restoring public trust in science. For example, encouraging post-publication peer review and increased transparency, are valuable tools for holding scientists and policymakers accountable. Additionally, social media platforms must take greater responsibility for the delicate balance of protecting freedom of speech while curbing the spread of misinformation.

## Conclusion

Our analysis of Iranian data from the COVID-19 pandemic shows how far science-centric populism can go when it comes to public health. Political populistic related decisions such as opposing foreign vaccines and pushing pseudoscientific contraceptives crippled Iran’s vaccine programme and caused thousands of preventable deaths. In our comparison of WHO data and The Economist’s excess deaths model, we found that in Iran, late vaccine introductions resulted in substantially greater excess mortality than in earlier vaccine-administered countries. Our analysis makes current events unfolding in the United States truly alarming.

The Iranian experience is the stark reminder that public health must be protected from ideological and populist overreach to forestall future crises. The global scientific community needs to increase public confidence through open debate and hold policymakers accountable. Only with collective action that we can protect public health and scientific integrity so that evidence-based policy trumps political rhetoric. As global scientists and researchers, we must engage in reflexive practice to preserve our integrity, hold ourselves to the highest scientific rigour, and engage with the lay public at every opportunity to promote science literacy and an understanding of the scientific method. Only in this way can public trust in scientific expertise be rebuilt, regardless of the decisions of individual political leaders.

## Data Availability

https://github.com/TheEconomist/covid-19-the-economist-global-excess-deaths-model
https://data.who.int/dashboards/covid19/data

https://data.who.int/dashboards/covid19/data

## Author Contributions

Nima Taefehshokr contributed to the literature search and preparation of the initial draft of the manuscript. Brielle Martens performed the analysis for Figures 1 and 2. Iman Beheshti used machine learning tools to prepare Figure 3 and interpreted the results. Rene P. Zahedi assited in developing the initial concept and drafted the PubPeer statement. Marek J. Los, an expert in COVID-19, provided valuable scientific insights and facilitated interactions with Iranian academics with respect to COVID-19 data. Jason Kindrachuk provided expert insights on COVID-19 transmission and pathogenesis and assisted in preparing the data analyses for Figures 1 and 2. Saeid Ghavami led the team, conceptualized the overall idea, engaged with Iranian scientists within Iran to interpret the analysis, and performed the final editing and refinement of the manuscript. All the authors have read and approved the manuscript in its final form.

## Data Availability

The data we have used are available through the references we have addressed in the article. All of the data are public data.

## Acknowledgements

The authors acknowledge Karen Limbert Rempel for professional English editing of the manuscript. The authors also acknowledge Mahan Ghaffari for scientific advice. Authors acknowledge the pre-print version of your article: (https://www.medrxiv.org/content/10.1101/2024.12.28.24319727v1)

## References

1. Hallin, D.C., et al., Pandemic communication in times of populism: Politicization and the COVID communication process in Brazil, Poland, Serbia and the United States. Soc Sci Med, 2024. 360: p. 117304.

2. Cordeiro-Rodrigues, L., D.L. Cole, and D. Duan, The Instrumentalization of Public Health Issues for Propaganda by the Far-Right. J Bioeth Inq, 2024.

3. Bonikowski, B., et al., Populism and nationalism in a comparative perspective: a scholarly exchange. Nations and Nationalism, 2019. 25(1): p. 58–81.

4. Mede, N.G., et al., Who supports science-related populism? A nationally representative survey on the prevalence and explanatory factors of populist attitudes toward science in Switzerland. PLoS One, 2022. 17(8): p. e0271204.

5. Taefehshokr, N., et al., The Threat of Science-Related Populism to Global Public Health: Lessons from Iran. medRxiv, 2024: p. 2024.12.28.24319727.

6. Nature. How Trump 2.0 is reshaping science. 31 March 2025 [cited 2025 April 3]; Available from: https://www.nature.com/collections/jcjhabjhgi.

7. Meyer, J.M., Power and Truth in Science-Related Populism: Rethinking the Role of Knowledge and Expertise in Climate Politics. Political Studies, 2024. 72(3): p. 845–861.

8. Wajner, D.F., S. Destradi, and M. Zürn, The effects of global populism: assessing the populist impact on international affairs. International Affairs, 2024. 100(5): p. 1819–1833.

9. Speed, E. and R. Mannion, Populism and health policy: three international case studies of right-wing populist policy frames. Sociol Health Illn, 2020. 42(8): p. 1967–1981.

10. Iran Physics Society. The opinion of the Iranian Physics Society on the construction of a device for detecting nanometric coronavirus using electromagnetic and remote methods. 2020 [cited 2024 7 October]; Available from: https://www.psi.ir/news2_fa.asp?id=3036.

11. Saliba, E. and L. Gharagozlou. Iran’s Revolutionary Guard Corps says its handheld device can detect coronavirus, scientists scoff. 2020 20 April [cited 2024 31 October]; Available from: https://www.nbcnews.com/news/world/iran-s-revolutionary-guard-corps-says-its-handheld-device-can-n1186346.

12. Von Hein, S., Taking advantage of Iran’s ‘patriotic’ vaccination policy, in DW News. 2021, Deutsche Welle: Gernamy

13. Retraction: “Safety and immunogenicity of a recombinant receptor-binding domain-based protein subunit vaccine (Noora vaccine) against COVID-19 in adults: A randomized, double-blind, placebo-controlled, Phase 1 trial”. J Med Virol, 2024. 96(3): p. e29509.

14. Dashti, N., et al., Comparative Immunogenicity and Neutralization Potency of Four Approved COVID-19 Vaccines in BALB/c Mice. Iran J Immunol, 2024. 21(1): p. 1–14.

15. GitHub, T.E. TheEconomist/covid-19-the-economist-global-excess-deaths-model: The Economist’s model to estimate excess deaths to the covid-19 pandemic. 2024 August 2024 [cited 2024 December 06].

16. Organization, W.H. World Health Organization. COVID-19 vaccines | WHO COVID-19 dashboard 2024 2024 [cited 2024 December 06]; Available from: https://data.who.int/dashboards/covid19/vaccines.

17. Organization, W.H. World Health Organization. COVID-19 data | WHO COVID-19 dashboard. 2023 [cited 2024 December 10]; Available from: https://data.who.int/dashboards/covid19/data.

18. Wickham H, et al. dplyr: A Grammar of Data Manipulation. 2023 [cited 2024 December 12]; Available from: https://dplyr.tidyverse.org.

19. G., G. and W. H, Dates and Times Made Easy with lubridate. Journal of Statistical Software 2011. 40: p. 1–25.

20. H, W. ggplot2: Elegant Graphics for Data Analysis 2016 [cited 2024 December 09]; Available from: https://ggplot2.tidyverse.org.

21. CO, W. cowplot: Streamlined Plot Theme and Plot Annotations for “ggplot2” 2024; Available from: https://wilkelab.org/cowplot/.

22. Ghafari, M., et al., A quantitative evaluation of the impact of vaccine roll-out rate and coverage on reducing deaths: insights from the first 2 years of COVID-19 epidemic in Iran. BMC Med, 2023. 21(1): p. 429.

23. A, K. rstatix: Pipe-Friendly Framework for Basic Statistical Tests. 2023 [cited 2024 December 09]; Available from: https://rpkgs.datanovia.com/rstatix/.

24. Watson, O.J., et al., Global impact of the first year of COVID-19 vaccination: a mathematical modelling study. Lancet Infect Dis, 2022. 22(9): p. 1293–1302.

25. Tadbiri, H., M. Moradi-Lakeh, and M. Naghavi, All-cause excess mortality and COVID-19-related deaths in Iran. Med J Islam Repub Iran, 2020. 34: p. 80.

26. Ghafari, M., A. Kadivar, and A. Katzourakis, Excess deaths associated with the Iranian COVID-19 epidemic: A province-level analysis. Int J Infect Dis, 2021. 107: p. 101–115.

27. Taylor, S.J. and B. Letham, Forecasting at Scale. American Statistician, 2018. 72(1): p. 37–45.

28. Bastani, M.N., et al., Comprehensive assessment of COVID-19 case fatality rate and influential factors in Khuzestan Province, Iran: a two-year study. J Health Popul Nutr, 2024. 43(1): p. 193.

29. Moradi-Lakeh, M. and A. Esteghamati, National Immunization Program in Iran: whys and why nots. Hum Vaccin Immunother, 2013. 9(1): p. 112–4.

30. Collaborators, G.B.D.I., Health system performance in Iran: a systematic analysis for the Global Burden of Disease Study 2019. Lancet, 2022. 399(10335): p. 1625–1645.

31. Duroseau, B., N. Kipshidze, and R.J. Limaye, The impact of delayed access to COVID-19 vaccines in low-and lower-middle-income countries. Front Public Health, 2022. 10: p. 1087138.

32. Van Egeren, D., et al., Vaccines Alone Cannot Slow the Evolution of SARS-CoV-2. Vaccines (Basel), 2023. 11(4).

33. Bolsen, T. and R. Palm, Politicization and COVID-19 vaccine resistance in the U.S. Prog Mol Biol Transl Sci, 2022. 188(1): p. 81–100.

34. Schenkel, M., Health emergencies, science contrarianism and populism: A scoping review. Soc Sci Med, 2024. 346: p. 116691.

35. Kozlov, M., Exclusive: NIH to cut grants for COVID research, documents reveal. Nature, 2025. 640(8057): p. 17–18.

